# Potential impact of annual vaccination with reformulated COVID-19 vaccines: lessons from the U.S. COVID-19 Scenario Modeling Hub

**DOI:** 10.1101/2023.10.26.23297581

**Authors:** Sung-mok Jung, Sara L. Loo, Emily Howerton, Lucie Contamin, Claire P. Smith, Erica C. Carcelén, Katie Yan, Samantha J. Bents, John Levander, Jessi Espino, Joseph C. Lemaitre, Koji Sato, Clif D. McKee, Alison L. Hill, Matteo Chinazzi, Jessica T. Davis, Kunpeng Mu, Alessandro Vespignani, Erik T. Rosenstrom, Sebastian A. Rodriguez-Cartes, Julie S. Ivy, Maria E. Mayorga, Julie L. Swann, Guido España, Sean Cavany, Sean M. Moore, Alex Perkins, Shi Chen, Rajib Paul, Daniel Janies, Jean-Claude Thill, Ajitesh Srivastava, Majd Al Aawar, Kaiming Bi, Shraddha Ramdas Bandekar, Anass Bouchnita, Spencer J. Fox, Lauren Ancel Meyers, Przemyslaw Porebski, Srini Venkatramanan, Aniruddha Adiga, Benjamin Hurt, Brian Klahn, Joseph Outten, Jiangzhuo Chen, Henning Mortveit, Amanda Wilson, Stefan Hoops, Parantapa Bhattacharya, Dustin Machi, Anil Vullikanti, Bryan Lewis, Madhav Marathe, Harry Hochheiser, Michael C. Runge, Katriona Shea, Shaun Truelove, Cécile Viboud, Justin Lessler

## Abstract

**Importance:** COVID-19 continues to cause significant hospitalizations and deaths in the United States. Its continued burden and the impact of annually reformulated vaccines remain unclear.

**Objective:** To project COVID-19 hospitalizations and deaths from April 2023–April 2025 under two plausible assumptions about immune escape (20% per year and 50% per year) and three possible CDC recommendations for the use of annually reformulated vaccines (no vaccine recommendation, vaccination for those aged 65+, vaccination for all eligible groups).

**Design:** The COVID-19 Scenario Modeling Hub solicited projections of COVID-19 hospitalization and deaths between April 15, 2023–April 15, 2025 under six scenarios representing the intersection of considered levels of immune escape and vaccination. State and national projections from eight modeling teams were ensembled to produce projections for each scenario.

**Setting:** The entire United States.

**Participants:** None.

**Exposure:** Annually reformulated vaccines assumed to be 65% effective against strains circulating on June 15 of each year and to become available on September 1. Age and state specific coverage in recommended groups was assumed to match that seen for the first (fall 2021) COVID-19 booster.

**Main outcomes and measures:** Ensemble estimates of weekly and cumulative COVID-19 hospitalizations and deaths. Expected relative and absolute reductions in hospitalizations and deaths due to vaccination over the projection period.

**Results:** From April 15, 2023–April 15, 2025, COVID-19 is projected to cause annual epidemics peaking November–January. In the most pessimistic scenario (high immune escape, no vaccination recommendation), we project 2.1 million (90% PI: 1,438,000–4,270,000) hospitalizations and 209,000 (90% PI: 139,000–461,000) deaths, exceeding pre-pandemic mortality of influenza and pneumonia. In high immune escape scenarios, vaccination of those aged 65+ results in 230,000 (95% CI: 104,000–355,000) fewer hospitalizations and 33,000 (95% CI: 12,000–54,000) fewer deaths, while vaccination of all eligible individuals results in 431,000 (95% CI: 264,000–598,000) fewer hospitalizations and 49,000 (95% CI: 29,000–69,000) fewer deaths.

**Conclusion and Relevance:** COVID-19 is projected to be a significant public health threat over the coming two years. Broad vaccination has the potential to substantially reduce the burden of this disease.

**Key points:** *Question:* What is the likely impact of COVID-19 from April 2023–April 2025 and to what extent can vaccination reduce hospitalizations and deaths?

*Findings:* Under plausible assumptions about viral evolution and waning immunity, COVID-19 will likely cause annual epidemics peaking in November–January over the two-year projection period. Though significant, hospitalizations and deaths are unlikely to reach levels seen in previous winters. The projected health impacts of COVID-19 are reduced by 10–20% through moderate use of reformulated vaccines.

*Meaning:* COVID-19 is projected to remain a significant public health threat. Annual vaccination can reduce morbidity, mortality, and strain on health systems.

## Introduction

Three and a half years after the SARS-CoV-2 virus first emerged in Wuhan, China, it seems the global community has transitioned from confronting COVID-19 as a pandemic emergency to managing it as an endemic, seasonally recurring virus [1]. While widespread immunity against SARS-CoV-2 has been achieved globally through vaccination and infections [2], the continued evolution of the virus causes antigenic changes and raises the potential for recurrent epidemics [3,4]. Current evidence suggests that both patterns of human contact and environmental factors contribute to seasonality in the intensity of SARS-CoV-2 transmission [5–7]. Combined, seasonality and ongoing “antigenic drift” of SARS-CoV-2 make it highly likely that the virus will pose a persistent threat to public health for the foreseeable future.

Going forward, one of the main tools for mitigating the impact of annual COVID-19 epidemics will be vaccination. As with influenza [8,9], continued antigenic drift of SARS-CoV-2 and intrinsic waning of the protection offered by previous vaccinations and infections (i.e., loss of immunity due to waning of immune protection, independent of the evolution of the virus) means regular re-vaccination with reformulated SARS-CoV-2 vaccines will be needed to mitigate the virus’s impact [10]. However, legitimate questions exist about how effective annual vaccination campaigns can be, given SARS-CoV-2’s rapid evolution, and what age ranges should be targeted, given the concentration of severe COVID-19 outcomes in older populations [11]. Hence, well-grounded projections of COVID-19’s impact under different vaccination scenarios help inform future vaccination policy.

The U.S. COVID-19 Scenario Modeling Hub (SMH) is a long-standing multi-team modeling effort that aims to project how the COVID-19 epidemic is likely to unfold in the mid- to long-term under various conditions [12,13]. These planning scenarios contrast various interventional strategies, characteristics of future viral variants, and other epidemiological or behavioral uncertainties, to provide projections of COVID-19 hospitalizations and deaths under each set of assumptions. By summarizing the results of multiple teams working on the same set of scenarios, the SMH takes advantage of the proven increased reliability of ensemble-based predictions over individual models [14]. Ensemble approaches have proven useful in multiple fields and across pathogens to inform public health policy, situational awareness, and individual decision-making [12].

Here, we present the results of applying the SMH approach to project the likely course of the COVID-19 epidemic in the United States over a two-year period (April 15, 2023–April 15, 2025) under different assumptions about the average speed of antigenic drift and possible recommendations for the use of reformulated annual COVID-19 vaccines from the Centers for Disease Control and Prevention (CDC).

## Methods

To estimate the potential impact of vaccination on COVID-19 hospitalizations and deaths, we invited multiple teams in an open call to provide two years of projections for six scenarios within the SMH framework [13,14]. Teams had broad discretion in the details of model implementation within scenario definitions (see below). Individual team projections were combined to produce ensemble projections for each scenario as well as an ensemble estimate of the expected impact of vaccination.

### Scenario definitions

Six scenarios were created representing the intersection of two axes: one representing the average speed of antigenic drift (i.e., immune escape) over the two-year projection period, and the second representing differing assumptions about CDC recommendations for, and uptake of, a reformulated SARS-CoV-2 vaccine. The antigenic drift axis consisted of (1) a ‘low immune escape’ scenario, where the SARS-CoV-2 virus evolves away from the immune signature of circulating variants at a rate of 20% per year (e.g., a vaccine with efficacy against symptomatic infection of 65% on June 15, 2023 is assumed to have an efficacy of 0.8✕0.65=52% one year later in the absence of immune waning), and (2) a ‘high immune escape’ scenario with an immune escape rate of 50% per year.

The vaccination axis consisted of three levels based on possible COVID-19 vaccine recommendations under consideration by the CDC Advisory Committee on Immunization Practices (ACIP): (1) no recommendation for annual vaccination with a reformulated vaccine, (2) a recommendation for those aged 65 and above (65+), and (3) a recommendation for all eligible groups. Across all scenarios, the vaccine is assumed to be reformulated to match the predominant variants circulating as of June 15 each year, and to become available to the public on September 1 of the same year. The annual uptake of reformulated vaccines in recommended groups is projected to follow the age group specific (0–17, 18–64, and 65+) uptake patterns observed for the first booster dose in each state (i.e., the first additional dose of vaccines after completing the primary series, authorized in September 2021) [15]. Uptake is assumed to saturate at levels reached one year after the recommendation (full uptake assumptions available on GitHub[16]; corresponding to 9% coverage in ages 0–17, 33% in 18–64, and 65% in 65+ nationally). Reformulated vaccines are presumed to have 65% vaccine effectiveness against symptomatic disease at the time of reformulation, with protection declining based on waning immunity and antigenic drift.

All contributing models were directed to incorporate waning immunity, with a requirement that the median waning time of protection against infection aligned with the designated range of 3–10 months. Furthermore, the incorporation of SARS-CoV-2 seasonality was required, though teams had discretion in terms of its implementation. Teams were directed not to consider changes in non-pharmaceutical interventions over the projection period. Full scenario details are available on GitHub[16].

### Ensemble projections

Eight different modeling teams contributed projections of weekly incident and cumulative COVID-19 hospitalizations and deaths for April 15, 2023–April 15, 2025 for all states and at the national level (one additional team provided projections for only North Carolina). Each team provided up to 100 representative epidemic trajectories for each scenario and outcome.

Trajectories were used to generate a probability distribution of incident outcomes each week. Distributions at each week were combined using the trimmed-linear opinion pool method (LOP) to create ensemble projections [14,17–19]. All reported numbers for incident and cumulative deaths and hospitalizations, and associated projection intervals (Pis), come from this ensemble. To estimate the expected impact of vaccination, the mean and variance in cumulative deaths and hospitalizations were calculated over the period of interest based on submitted trajectories. Within each model, the expected impact of vaccination was determined by calculating the difference, or ratio, of projected deaths and hospitalizations between different vaccination scenarios sharing the same rate of immune escape, with variances estimated using the Delta method [20]. These individual model level estimates were then combined to produce an ensembled estimate of expected vaccine impact and associated confidence intervals (CIs) using standard meta-analysis techniques as implemented in the R package ‘*metafor*’ [21,22]. We note that in estimating vaccine impact we (1) take the vaccine impacts estimated by each model and then ensemble those (rather than looking at the impact in ensemble estimates) and (2) use different techniques in combining vaccine impact estimates aimed at getting expected values and confidence intervals (rather than predictions intervals). Hence, vaccine impact estimates are not directly reproducible by comparing ensemble trajectories (which are not mathematically equivalent).

## Results

Based on the ensemble of projections from eight contributing models under plausible assumptions about the viral evaluation and annual vaccination recommendations from the CDC, we project that between April 15, 2023 and April 15, 2025, the United States will experience annual COVID-19 epidemics peaking between November and January and causing approximately 1 million cumulative hospitalizations and 100,000 cumulative deaths each year (**Figure-1** & **Table-1**). The extent of COVID-19 impact over this period varies significantly by scenario, with 1.4 million (90% PI: 983,000–1,947,000) hospitalizations and 130,000 (90% PI: 71,000–201,000) deaths over the two-year projection period in the most optimistic scenario (reformulated vaccines recommended for all individuals, 20% immune escape) and 2.1 million (90% PI: 1,438,000–4,270,000) hospitalizations and 209,000 (90% PI: 139,000–461,000) deaths in the most pessimistic scenario (no recommendation, 50% immune escape) (**Figure-S1**). While significant, even in the most pessimistic scenario we project deaths and hospitalizations are unlikely to be as high as the peak weekly hospitalizations seen in the first Omicron wave in early 2022 (150,000 hospitalizations per week). Furthermore, projected weekly hospitalizations are likely to remain at or below CDC-designated medium community transmission levels (10–19 weekly hospitalizations per year)[23] across all scenarios (**Figure-1**). There is moderate variation between states in peak timing and size of COVID-19 epidemic waves, although most generally follow national trends (**Figures-S2 & S3**).

**Figure 1.**
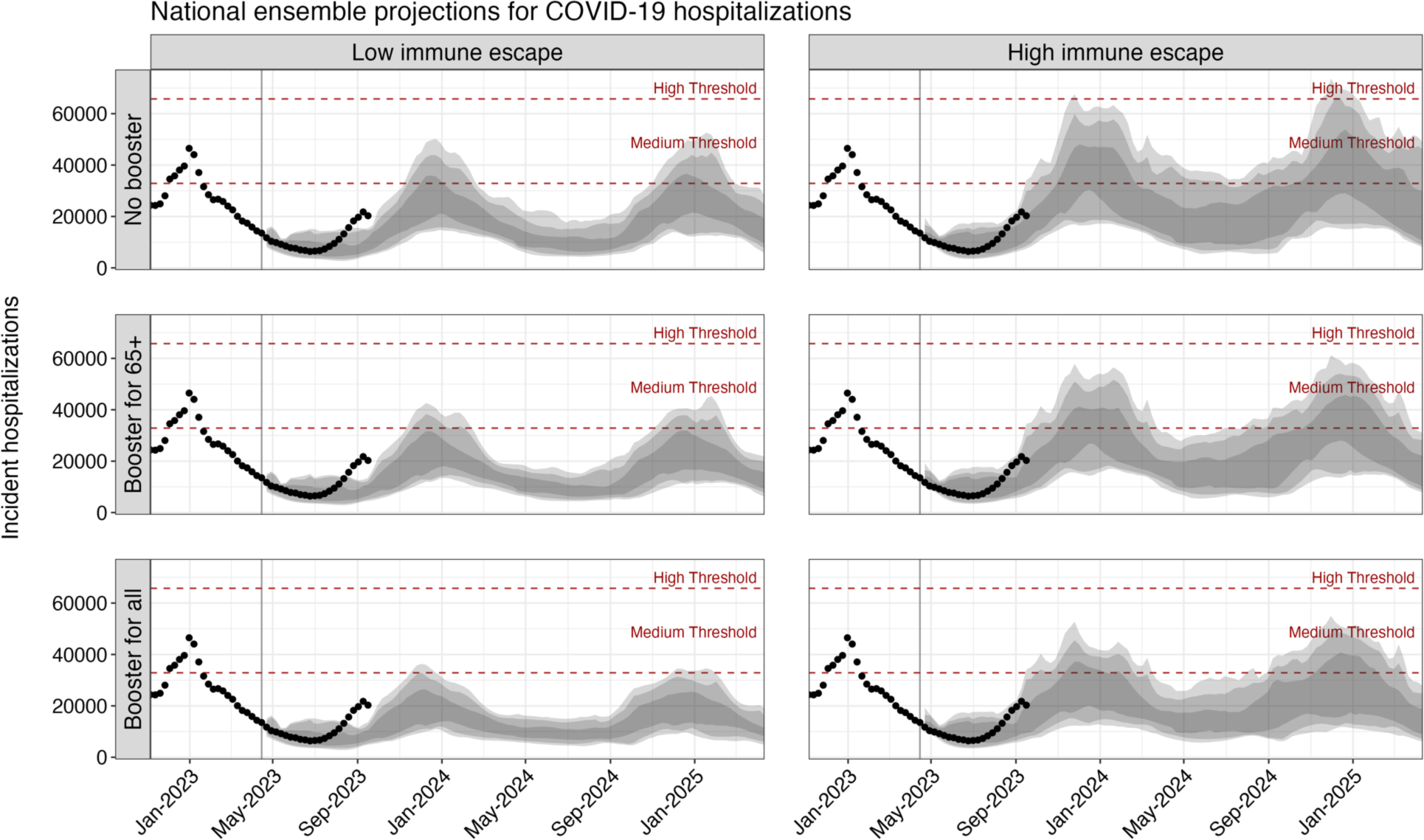
Projected weekly COVID-19 hospitalizations in the United States across scenarios, April 2023–April 2025. Ensemble projections from the COVID-19 Scenario Modeling Hub of national COVID-19 hospitalization for the period April 2023–April 2025 are shown by scenario. Dots indicate the observed weekly hospitalizations between December 2022 and August 12, 2023. Shading from lightest to darkest represents 90%, 80%, and 50% projection intervals. Red dashed lines correspond to the CDC-designated COVID-19 community-level indicators: medium (10–19 weekly hospitalizations per 100,000) and high (>20 weekly hospitalizations per 100,000) levels. The vertical line on April 15, 2023 marks the start of the projection period.

**Table 1.** Projected national peak timing and peak size of hospitalizations across scenarios.

|  | April 15, 2023–April 14, 2024 |  |  |  | April 15, 2024–April 15, 2025 |  |  |  |
| --- | --- | --- | --- | --- | --- | --- | --- | --- |
| Scenario | Peak timing | Peak size | Total hospitalizations | Total deaths | Peak timing | Peak size | Total hospitalizations | Total deaths |
| **High immune escape** |  |  |  |  |  |  |  |  |
| **No booster recommendation** | Dec 10<br>(Oct 15–Apr 14) | 42,000<br>(18,000–105,000) | 1,017,000<br>(767,000–2,058,000) | 100,000<br>(68,000–217,000) | Dec 15<br>(Oct 13–Apr 13) | 45,000<br>(17,000–90,000) | 1,093,000<br>(670,000–2,211,000) | 108,000<br>(71,000–244,000) |
| **Booster recommended for 65+** | Dec 10<br>(Oct 15–Feb 7) | 39,000<br>(17,000–91,000) | 943,000<br>(689,000–1,859,000) | 94,000<br>(55,000–178,000) | Dec 15<br>(Oct 13–Feb 23) | 41,000<br>(16,000–77,000) | 1,049,000<br>(584,000–1,959,000) | 99,000<br>(67,000–189,000) |
| **Booster recommended for all** | Dec 10<br>(Oct 8–Feb 18) | 35,000<br>(15,000–91,000) | 836,000<br>(595,000–1,723,000) | 82,000<br>(53,000–173,000) | Dec 8<br>(Jun 9–Feb 19) | 32,000<br>(14,000–77,000) | 949,000<br>(606,000–1,741,000) | 89,000<br>(64,000–182,000) |
| **Low immune escape** |  |  |  |  |  |  |  |  |
| **No booster recommendation** | Dec 13<br>(Aug 13–Apr 14) | 36,000<br>(16,000–81,000) | 825,000<br>(676,000–1,169,000) | 79,000<br>(57,000–124,000) | Dec 29<br>(Oct 27–Apr 13) | 35,000<br>(14,000–76,000) | 956,000<br>(578,000–1,304,000) | 85,000<br>(49,000–166,000) |
| **Booster recommended for 65+** | Dec 10<br>(Aug 13–Feb 18) | 34,000<br>(15,000–68,000) | 767,000<br>(620,000–1,020,000) | 70,000<br>(45,000–111,000) | Dec 22<br>(Oct 27–Mar 9) | 32,000<br>(13,000–65,000) | 857,000<br>(485,000–1,128,000) | 80,000<br>(34,000–109,000) |
| **Booster recommended for all** | Dec 3<br>(Apr 30–Mar 3) | 26,000<br>(13,000–57,000) | 670,000<br>(487,000–920,000) | 63,000<br>(38,000–101,000) | Dec 15<br>(Jun 12–Mar 9) | 28,000<br>(12,000–51,000) | 717,000<br>(496,000–1,027,000) | 67,000<br>(33,000–100,000) |

Each value represents the median projection with 90% projection interval below.

Ensemble projections indicate that annual vaccination has the potential to substantially reduce both hospitalizations and deaths from COVID-19 (**Figure-2**). In high immune escape scenarios, if vaccination is confined to 65+, and uptake patterns mirror what was seen for the first booster dose, we would expect a reduction in hospitalizations of 8% (95% CI: 5–12) compared to the no vaccination scenario and a reduction in deaths of 13% (95% CI: 7–18). This corresponds to absolute reductions of 230,000 (95% CI: 104,000–355,000) hospitalizations and 33,000 (95% CI: 12,000–54,000) deaths across the entire United States over the two-year projection period.

**Figure 2.**
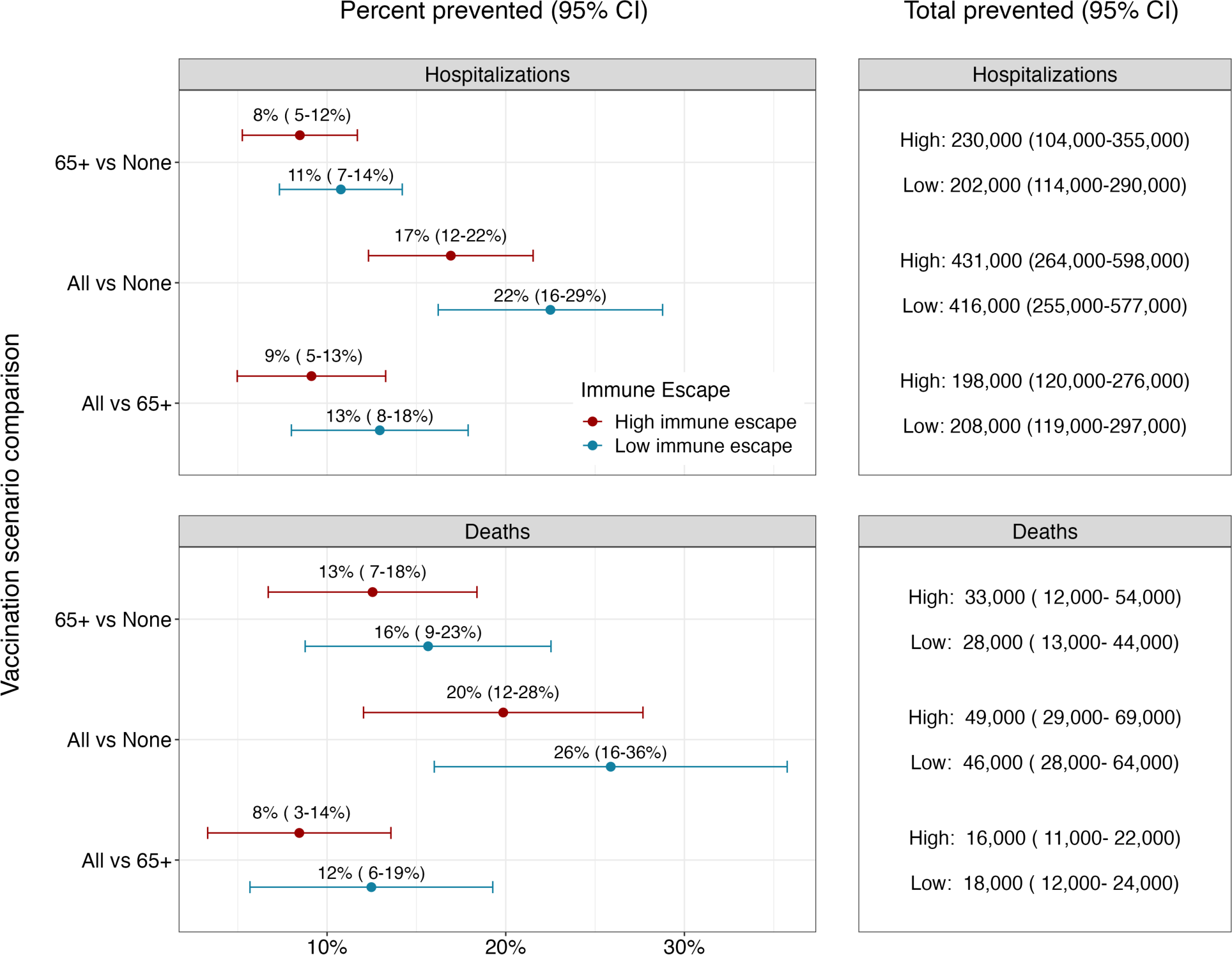
Percent and total prevented COVID-19 hospitalizations and deaths by annual vaccination recommendation with reformulated vaccines. Relative and absolute differences in cumulative hospitalizations and deaths over the next two years (April 2023–April 2025) between different vaccination recommendations. Red and blue dots and error bars represent the median and 95% confidence interval of percent prevented outcomes in high and low immune escape scenarios (50% per year and 20% per year), respectively.

Expanding vaccination recommendations to all individuals would lead to substantial additional reductions in deaths and hospitalizations (**Figure-2**). Under the assumption that coverage equivalent to the first booster dose is attained, vaccination of all individuals reduces hospitalizations by 9% (95% CI: 5–13, N = 198,000, 95% CI: 120,000–276,000) and deaths by 8% (95% CI: 3–14, N = 16,000, 95% CI: 11,000–22,000) compared to vaccination of 65+ alone in high immune escape scenarios. This corresponds to a total reduction of 17% (95% CI: 12–22, N = 431,000, 95% CI: 264,000–598,000) in hospitalizations and 20% (95% CI: 12–28, N = 49,000, 95% CI: 29,000–69,000) in deaths compared to the no vaccination scenario. Results are similar in low immune escape scenarios.

A significant factor contributing to state-level variation in the projected impact of vaccine recommendations is the assumed uptake level of reformulated vaccines (**Figure-3, S4, and S5**). States with higher coverage among 65+ are anticipated to experience substantial reductions in hospitalizations, exceeding 150 per 100,000 in high immune escape scenarios, if the reformulated vaccines are recommended to all. In contrast, the state with the lowest coverage in 65+, North Carolina, is expected to witness reductions of less than 75 per 100,000.

**Figure 3.**
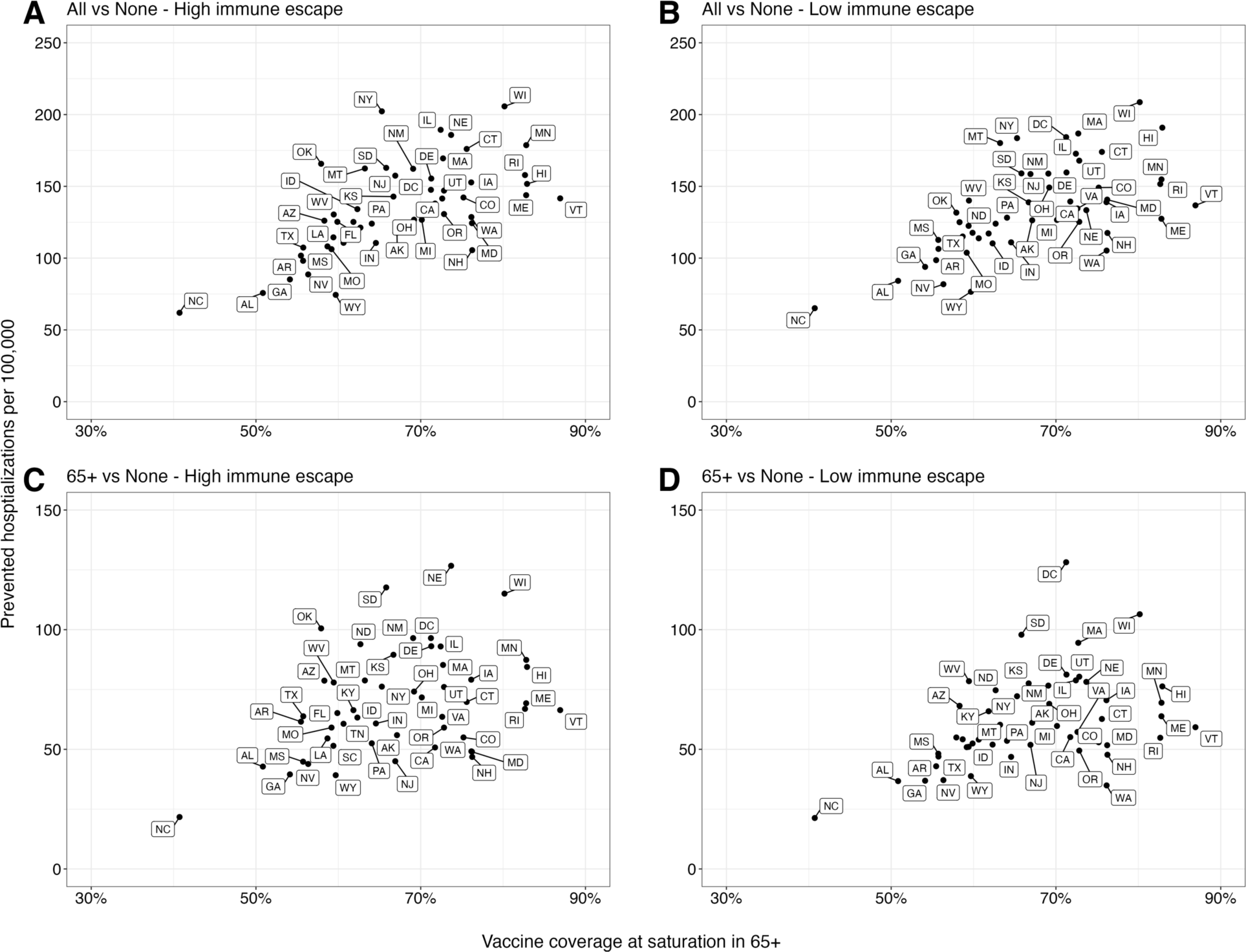
Relationship between prevented COVID-19 hospitalizations and assumed vaccine coverage in individuals aged 65 and above across US states. The relationship between the cumulative difference in COVID-19 hospitalizations for the next two years (April 2023–April 2025) under different vaccination recommendations and assumed vaccine uptake among those aged 65 and above (65+) in each US state: **(A & B)** vaccination of all compared to no vaccination, and **(C & D)** vaccination of 65+, compared to no vaccination. The x-axis represents the assumed vaccine coverage among 65+ at saturation considering the higher severity in 65+, and dots in each panel correspond to individual US states.

## Discussion

Based on the ensemble of projections from eight modeling teams for the next two years (April 2023–April 2025), it is expected that COVID-19 will remain a persistent public health threat in the United States for the foreseeable future. Nevertheless, our projections highlight that annual vaccination with reformulated vaccines can substantially mitigate this burden if coverage reaches levels observed for the first (i.e., fall 2021) COVID-19 booster.

Across all scenarios, our projections indicate that COVID-19 hospitalizations and deaths would be substantially less than what was seen in the early stages of the pandemic (e.g., between April 2021–April 2023 there were 4.2 million hospitalizations and 570,000 deaths [24]). Nonetheless, COVID-19 is projected to remain one of the leading causes of death in the United States [25]. For context, in our most pessimistic scenario (no CDC vaccine recommendation, high immune escape), annual COVID-19 mortality is expected to be similar to pre-pandemic mortality from Alzheimer’s disease (**Figure-4**), while in the most optimistic scenario (vaccines recommended for all, low immune escape) mortality would be similar to that seen from diabetes in the pre-pandemic period. In all cases, COVID-19 mortality is projected to exceed that of influenza and pneumonia.

**Figure 4:**
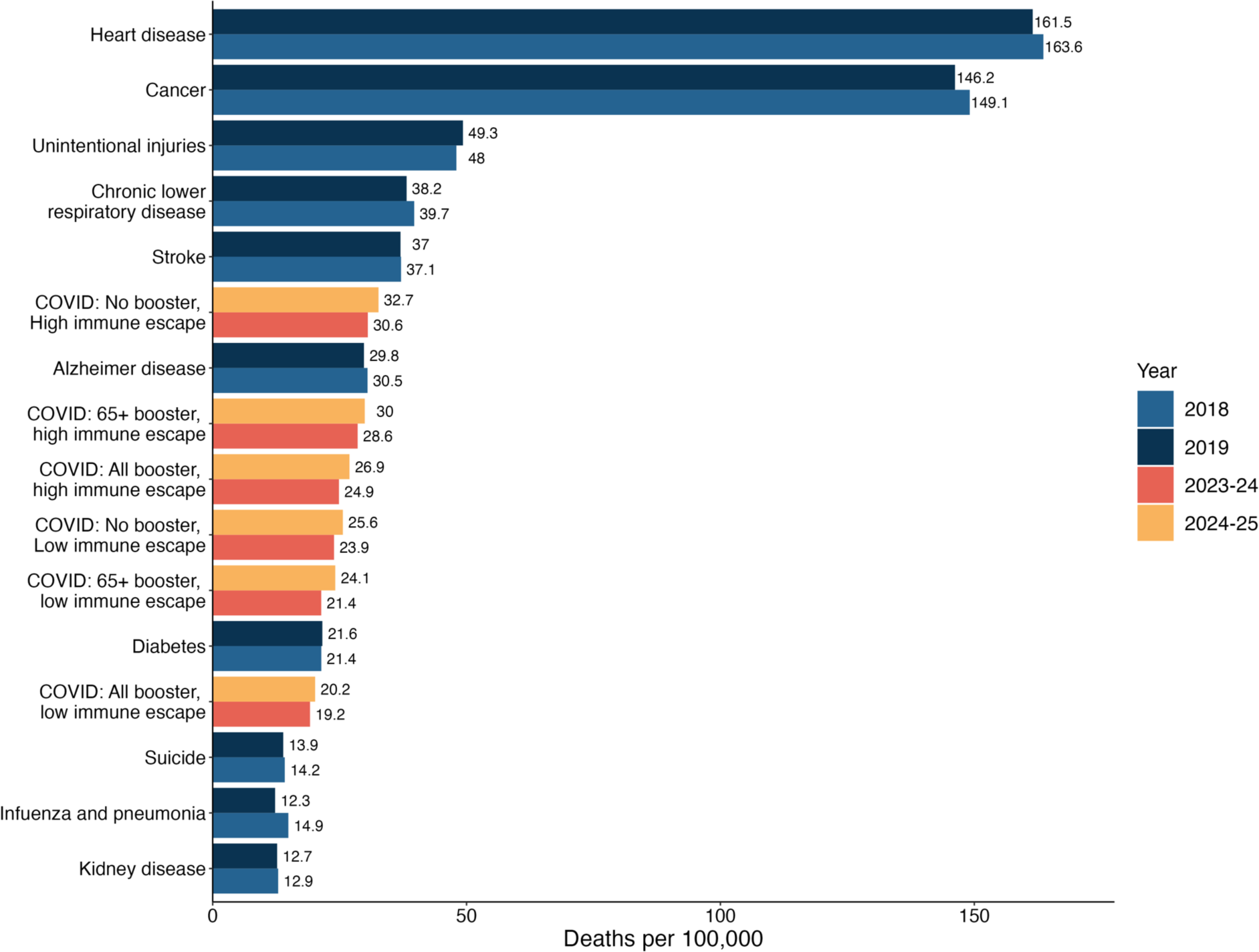
Comparison between the projected COVID-19 mortality by scenario and the 10 leading causes of pre-pandemic mortality in the United States. Projected COVID-19 mortality by scenario and by period (April 2023–April 2024 and April 2024–April 2025) are compared with the 10 leading causes of mortality in the United States, which were obtained from the CDC age-adjusted disease burden rates in the pre-pandemic period [25].

While the projected impact of annual vaccination on disease burden is significant, it is highly dependent on assumed vaccine uptake. This gives us reason for both caution and hope. Previous CDC booster recommendations, including that for the 2022 reformulated vaccine (i.e., bivalent vaccines authorized in August 2022), have not achieved the coverage observed for the first booster [26]. Reduced coverage would substantially blunt the impact of any vaccine recommendations. However, it is worth noting that many states where we assume low vaccination coverage, such as North Carolina and Pennsylvania, have not historically been ranked among the states with the lowest vaccine coverage for annual influenza vaccines [27], suggesting potential for increasing vaccine uptake in these regions.

As with any attempt to project into the future, our projections come with major caveats and limitations. First and foremost, scenario projections are conditional on often strict scenario assumptions. Both vaccine coverage and effectiveness might deviate considerably from scenario assumptions, although historical trends of influenza vaccination suggest that achieving higher coverage is unlikely (CDC, 2022b). Additionally, if future variants differ in intrinsic transmissibility or disease severity from that of the current Omicron lineages, the projected disease burden may alter accordingly. Furthermore, all scenarios were built on the assumption of continuous immune escape with a constant rate. However, the emergence of new SARS-CoV-2 variants showing a significant level of antigenic drift within a very short span (e.g., Omicron [29,30]) could increase the disease burden far beyond these projections.

## Conclusion

Despite its limitations, ensembling scenario-based projections from multiple teams is one of the most robust approaches for estimating COVID-19’s future burden and the potential benefits of vaccination, providing valuable information for public health planning. Our results show that COVID-19 will likely remain a major threat to human health in the United States in the coming years. In the face of this threat, broad vaccination against SARS-CoV-2 has the potential to save tens of thousands of lives each year.

### Data availability

All source data are openly available on COVID-19 Scenario Modeling Hub GitHub: https://github.com/midas-network/covid19-scenario-modeling-hub. Replication codes are available on GitHub: https://github.com/SungmokJung/covid19-scenario-modeling-hub-R1. Any use of trade, firm, or product names is for descriptive purposes only and does not imply endorsement by the U.S. Government.

### Author contributions

J. Lessler had full access to all of the data in the study and takes responsibility for the integrity of the data and the accuracy of the data analysis; Concept and design: J. Lessler, C. Viboud, K. Shea, S. Truelove, H. Hochheiser, M. Runge; Acquisition, analysis, or interpretation of data: All authors; Drafting of the manuscript: S. Jung, J. Lessler; Statistical analysis: S. Jung, S. Loo. E. Howerton, L. Contamin, S. Truelove, J. Lessler; Obtained funding: J. Lessler, S. Truelove, K. Shea, H. Hochheiser, A. Vespignani, J. Swann, S. Moore, L. Meyers, B. Lewis, M. Marathe; Administrative, technical, or material support: None; Supervision: J. Lessler, C. Viboud, K. Shea, S. Truelove, H. Hochheiser, M. Runge, A. Vespignani, J. Swann, S. Chen, S. Moore, A. Srivastava, L. Meyers, B. Lewis, M. Marathe.

### Conflict of Interest Disclosures

J. Espino is president of General Biodefense LLC, a private consulting group for public health informatics, and has interest in READE.ai, a medical artificial intelligence solutions company. M. Runge reports stock ownership in Becton Dickinson & Co., which manufactures medical equipment used in COVID-19 testing, vaccination, and treatment. J. Lessler has served as an expert witness on cases where the likely length of the pandemic was of issue.

### Funding/Support

S. Jung, S. Loo, C. Smith, E. Carcelén, J. Lemaitre, K. Sato, C. Mckee, S. Truelove, A. Hill, and J. Lessler were supported by Centers for Disease Control and Prevention (200-2016-91781). C. Smith, S. Truelove, and A. Hill were supported by the National Science Foundation (2127976). C. Smith, A. Hill, S. Truelove, and J. Lessler were supported by the US Department of Health and Human Services; Department of Homeland Security; California Department of Public Health; Johns Hopkins University. J. Lemaitre, C. Smith, A. Hill, S. Truelove, and J. Lessler were supported by Amazon Web Services. J. Lessler (R01GM140564) and J. Lemaitre (5R01AI102939) were supported by the National Institutes of Health. J. Lemaitre was supported by the Swiss National Science Foundation (200021-172578). L. Contamin, J. Levander, J. Espino, and H. Hochheiser were supported by NIGMS 5U24GM132013. E. Howerton and K. Shea were supported by NSF RAPID awards DEB-2028301, DEB-2037885, DEB-2126278, and DEB-2220903. K. Yan was supported by NSF Grant No. DGE1255832. E. Howerton was supported by the Eberly College of Science Barbara McClintock Science Achievement Graduate Scholarship in Biology at the Pennsylvania State University. M. Chinazzi, J. T. Davis, K. Mu, and A. Vespignani were supported by HHS/CDC 6U01IP001137, HHS/CDC 5U01IP0001137, and the Cooperative Agreement no. NU38OT000297 from the Council of State and Territorial Epidemiologists (CSTE). E. Rosenstrom, J. Ivy, M. Mayorga, and J. Swann were supported by TRACS/NIH grant UL1TR002489; CSTE and CDC cooperative agreement no. NU38OT000297. G. España and S. Moore were supported by Scenario Modeling Hub Consortium fellowship. S. Moore was supported by NIAID R21AI164391. A. Perkins was supported by NIGMS R35 MIRA program R35GM143029. K. Bi, S. Bandekar, A. Bouchnita, S. Fox, and L. Meyers were supported by CSTE NU38OT000297, CDC Supplement U01IP001136-Suppl, CDC 75D30122C14776 and NIH Supplement R01AI151176-Suppl. P. Porebski, S. Venkatramanan, A. Adiga, B. Lewis, B. Klahn, J. Outten, B. Hurt, H. Mortveit, A. Wilson, M. Marathe, J. Chen, S. Hoops, P. Bhattacharya, D. Machi acknowledge support from SMC Fellowship 75D30121F00005-2005604290, VDH Grant PV-BII VDH COVID-19 Modeling Program VDH-21-501-0135, NSF Grant No. OAC-1916805, NSF Expeditions in Computing Grant CCF-1918656, DTRA subcontract/ARA S-D00189-15-TO-01-UVA, and UVA strategic funds. Model computation was supported by NSF ACCESS CIS230005 and UVA. The funder had no role in the design and conduct of the study; collection, management, analysis, and interpretation of the data; preparation, review, or approval of the manuscript; and decision to submit the manuscript for publication.

## Supplementary figures

**Figure S1:**
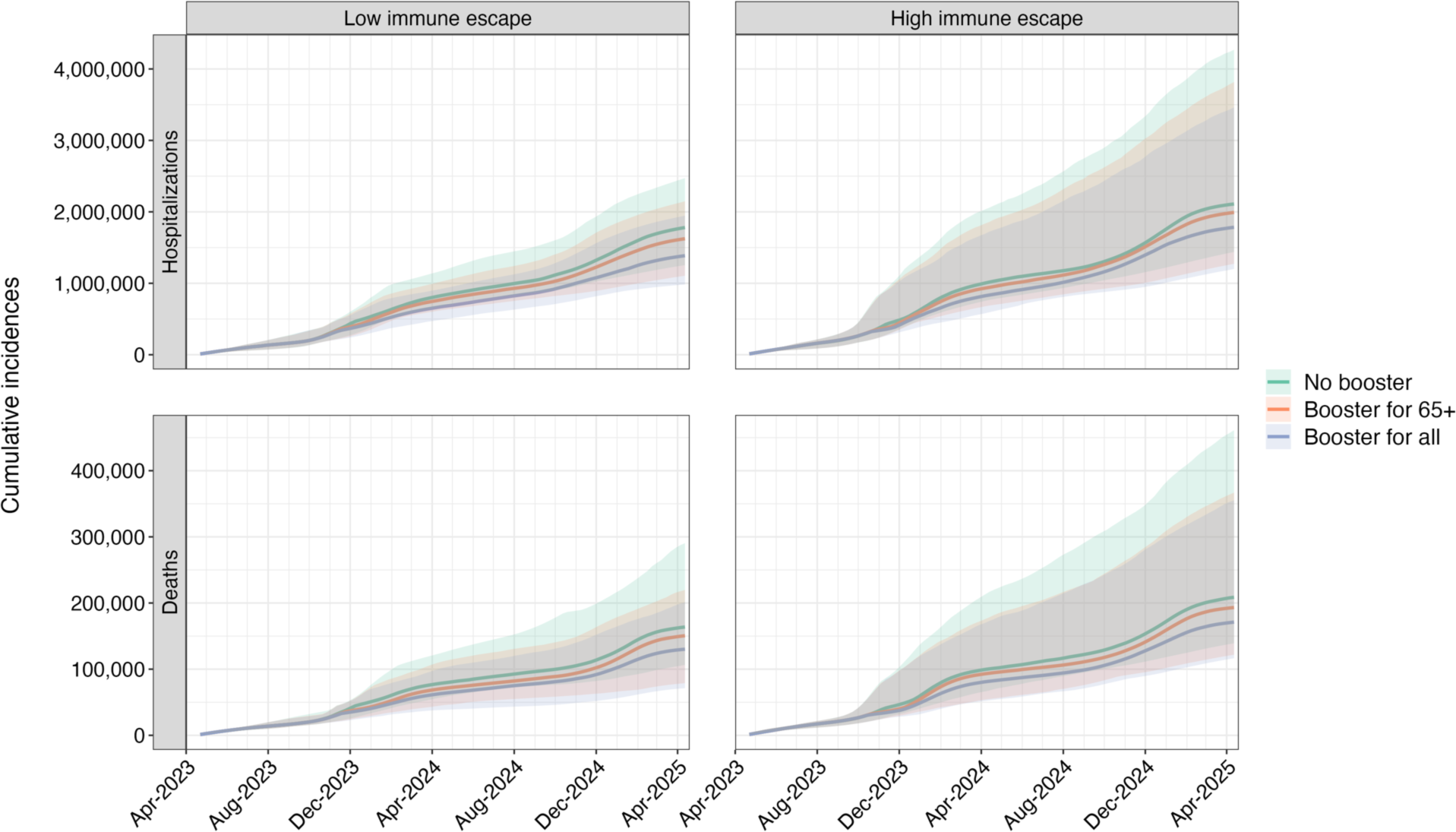
Projected cumulative COVID-19 hospitalizations and deaths in the United States by scenario, April 2023–April 2025. Ensemble projections for cumulative COVID-19 hospitalization and deaths in the United States for the next two years (April 2023–April 2025) are shown by scenario. Lines and shades indicate the median of projected outcomes and 90% projection intervals. Each color represents different annual vaccination recommendations (no recommendation, reformulated vaccines recommended for those aged 65 and above, and recommended for all age groups).

**Figure S2:**
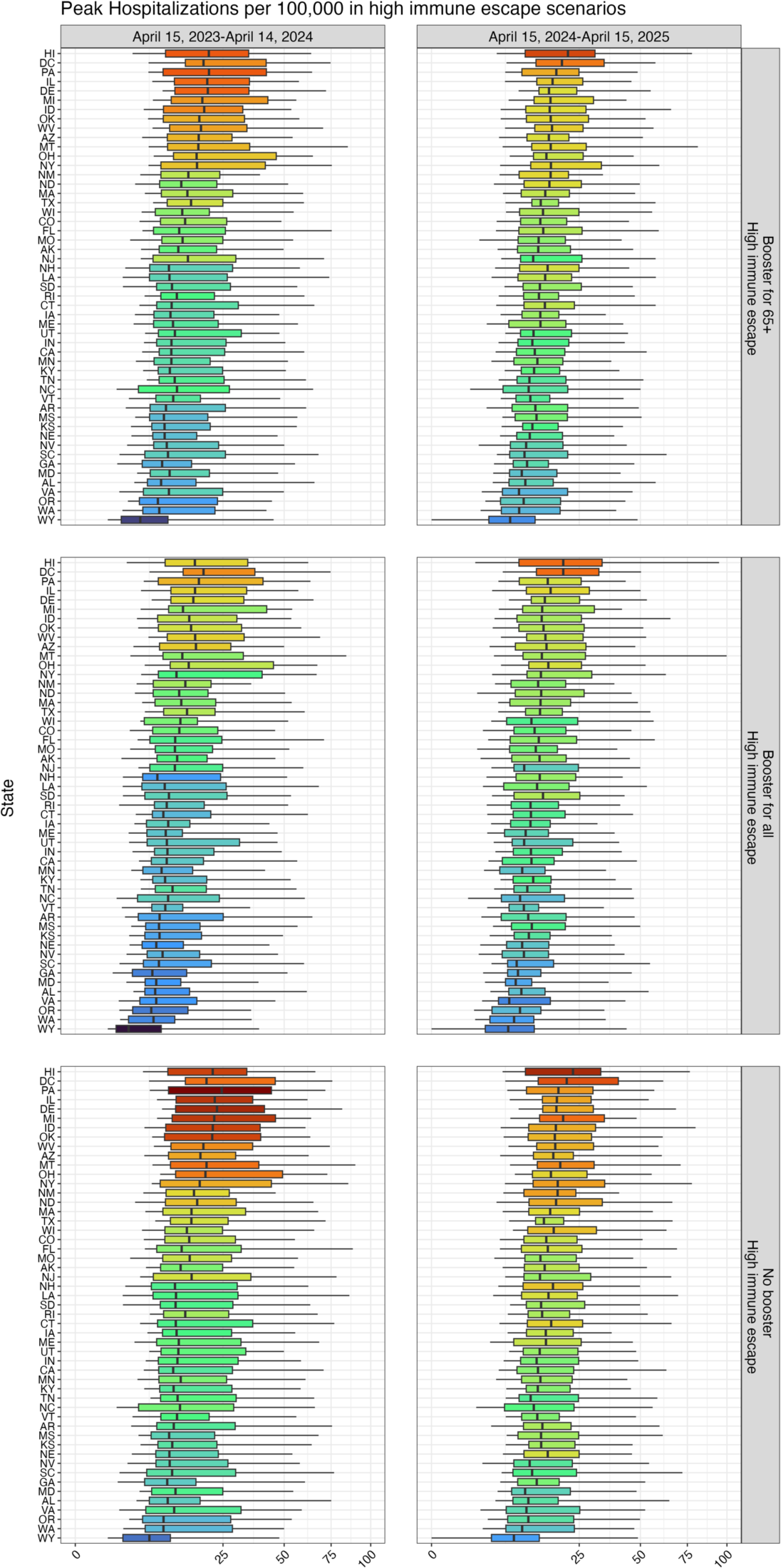
State-level peak COVID-19 hospitalizations in high immune escape scenarios by season and vaccination scenario. The peak hospitalizations per 100,000 over the next two years (April 2023–April 2025) under high immune escape assumption are shown by US state and by vaccination scenario (no recommendation, reformulated vaccines recommended for those aged 65 and above, and recommended for all age groups). Shades of blue indicate states with lower values and shades of red indicate states with higher values. For visualizations, square root scaling was applied in x-axes.

**Figure S3:**
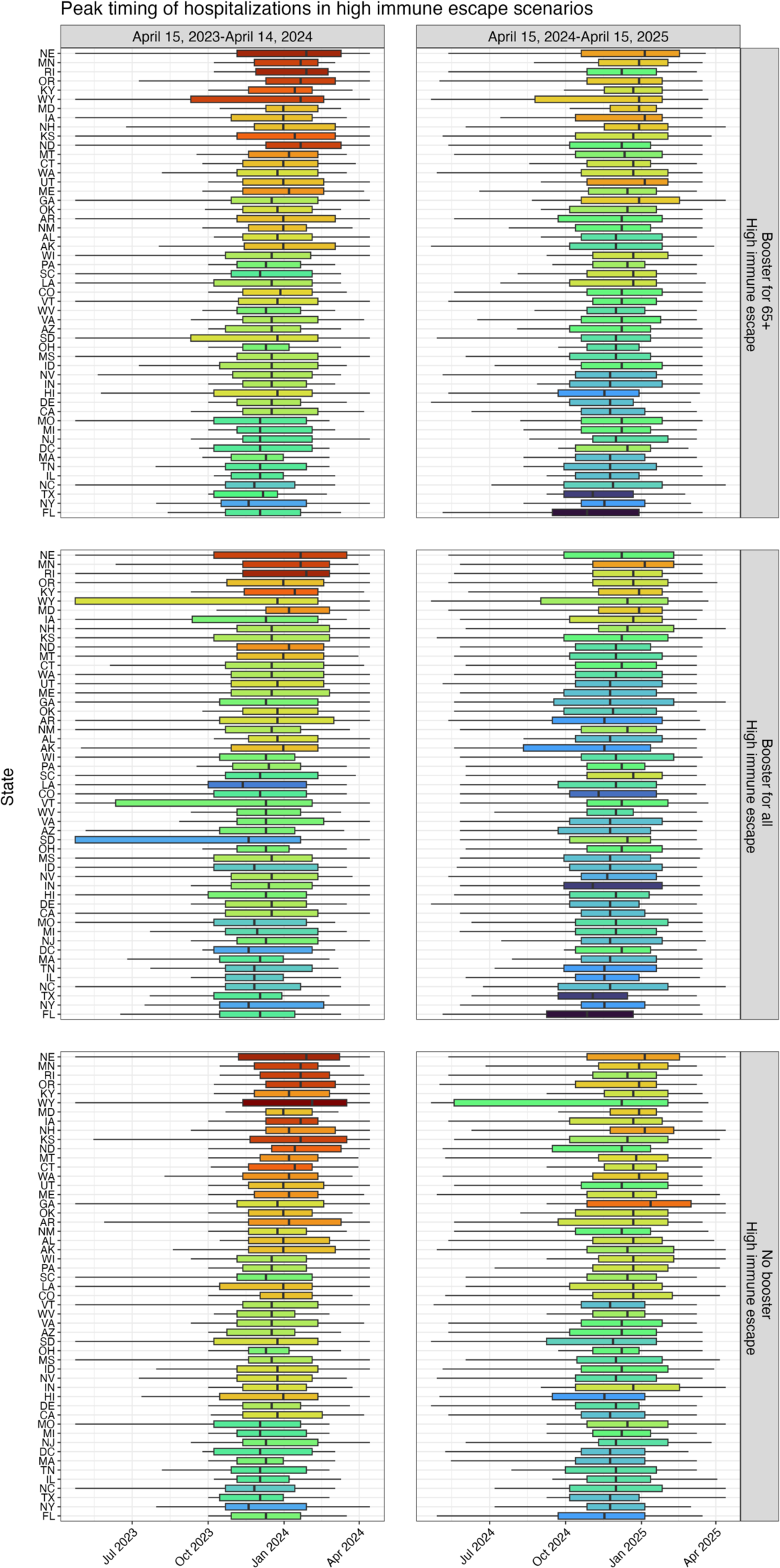
State-level peak timing of COVID-19 hospitalizations in high immune escape scenarios by season and vaccination scenario. The peak timing of hospitalizations under high immune escape assumption is shown by US state and by vaccination scenario (no recommendation, reformulated vaccines recommended for those aged 65 and above, and recommended for all age groups). Shades of blue indicate states with lower values and shades of red indicate states with higher values.

**Figure S4:**
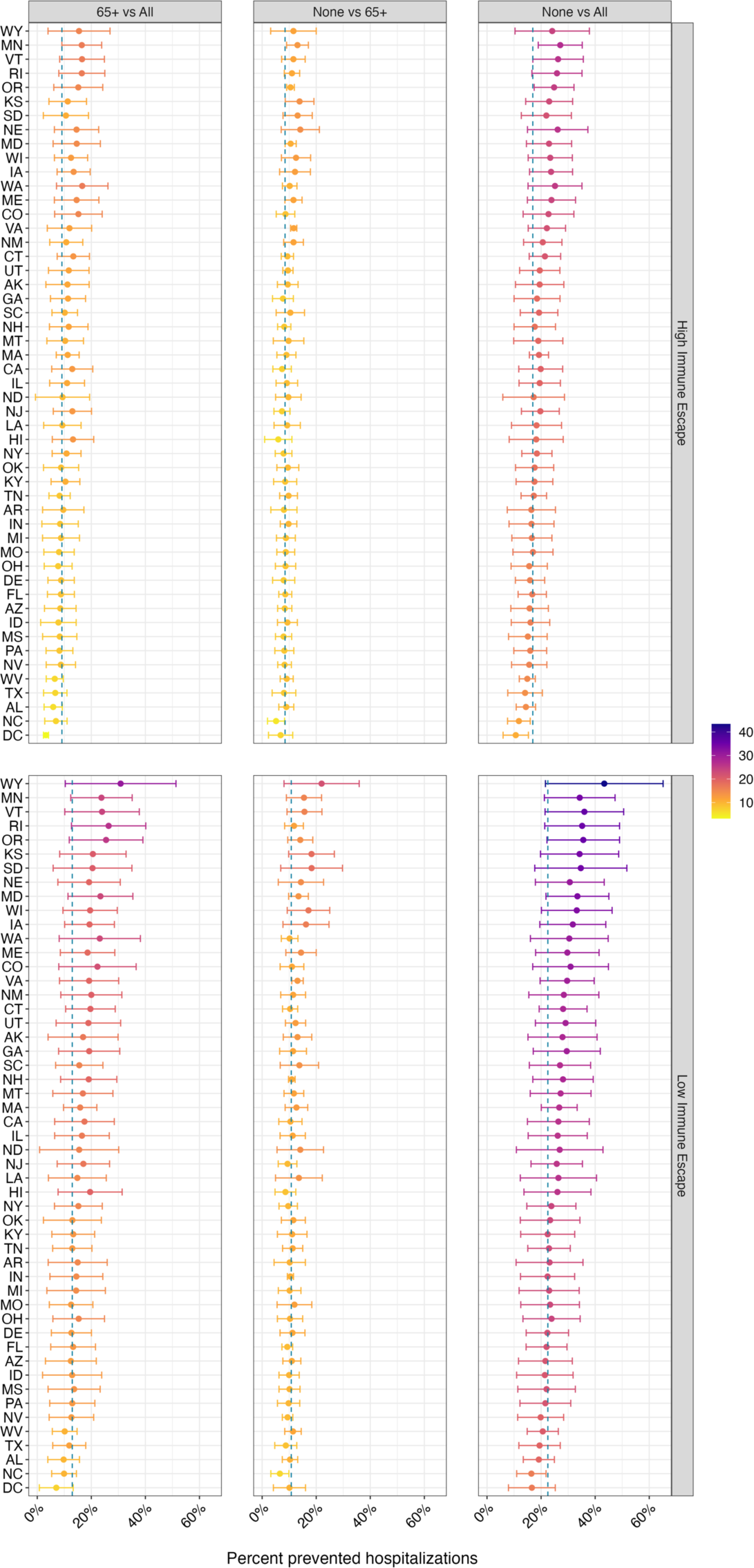
State-level percent prevented COVID-19 hospitalizations between the annual vaccination scenarios from April 2023 to April 2025 by scenario. Relative differences in cumulative COVID-19 hospitalizations over the next two years (April 2023–April 2025) between different vaccination scenarios are shown by immune escape level and by US state. Shades of yellow indicate states with lower values and shades of purple indicate states with higher values.

**Figure S5:**
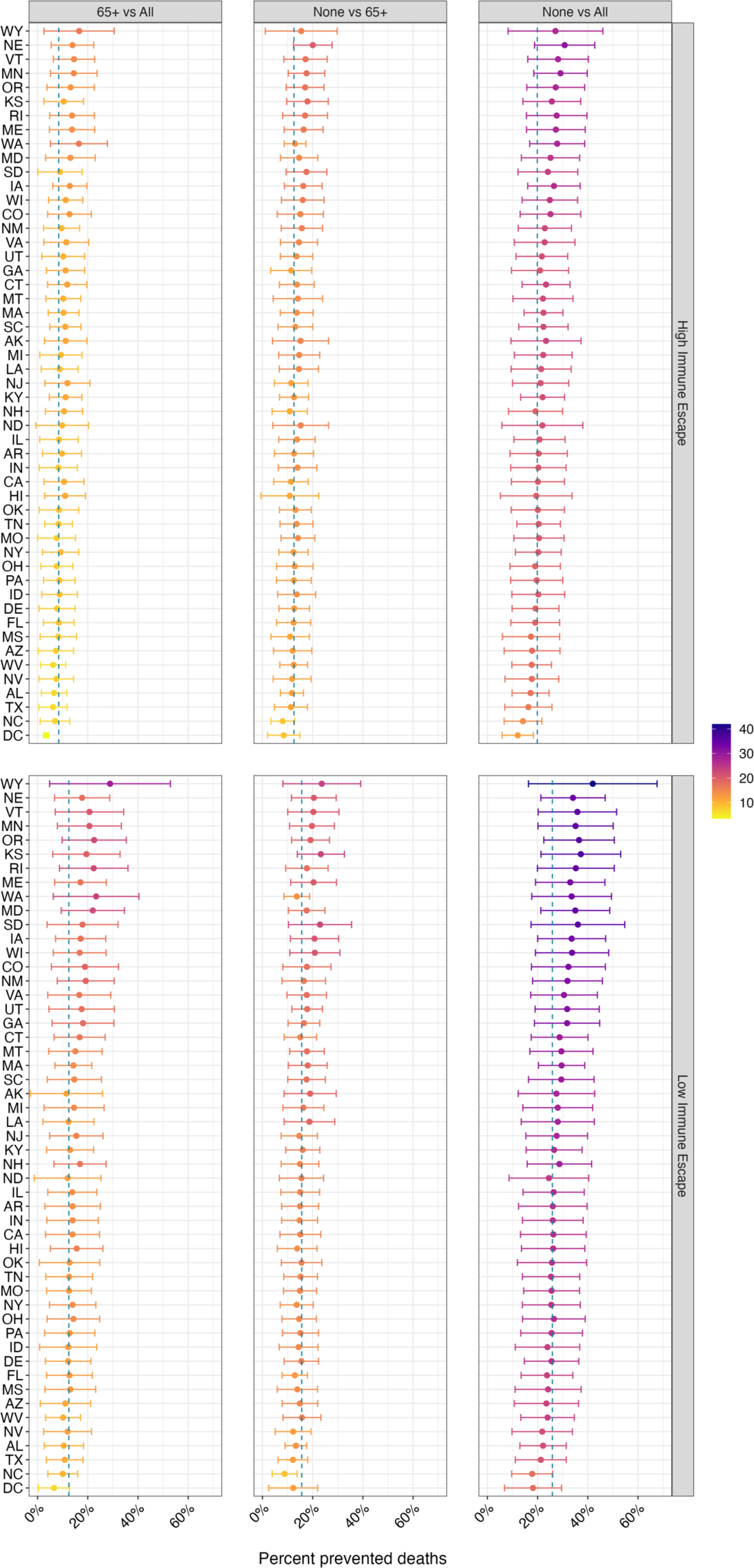
State-level percent prevented COVID-19 deaths between the annual vaccination scenarios from April 2023 to April 2025 by scenario. Relative differences in cumulative COVID-19 deaths over the next two years (April 2023–April 2025) between different vaccination scenarios are shown by immune escape level and by US state. Shades of yellow indicate states with lower values and shades of purple indicate states with higher values.

## Notes

### Summary of Updates

The author list was revised.

